# Associations between Affect, Physical Activity, and Anxiety Among US Children During COVID-19

**DOI:** 10.1101/2020.10.20.20216424

**Authors:** Jasmin M. Alves, Alexandra G. Yunker, Alexis DeFendis, Anny H. Xiang, Kathleen A. Page

**Affiliations:** Division of Endocrinology, Department of Medicine, Keck School of Medicine, University of Southern California, Los Angeles, CA 90089; Diabetes and Obesity Research Institute, Keck School of Medicine, University of Southern California, Los Angeles CA 90089; Department of Research and Evaluation, Kaiser Permanente Southern California, Pasadena, CA 91101

**Keywords:** COVID-19, Affect, State Anxiety, Physical Activity, Leisure Screen Time, US children

## Abstract

We investigated how emotional responses (positive and negative affect), physical activity (PA), and sedentary behaviors related to anxiety among US children during the COVID-19 pandemic. Sixty-four typically-developing children (63% girls) age 9-15 years old completed two virtual visits during height of “stay-at-home” measures between April 22 – July 29, 2020. Children completed 24-hour PA recalls, state portion of State-Trait Anxiety Inventory for Children (STAIC), and the shortened 10-item Positive and Negative Affect Schedule for Children (PANAS-C). Children reported state anxiety scores that were more than 5 standard deviations greater than values from healthy pediatric populations prior to the pandemic. Children with higher positive affect and who reported more time in PA reported less state anxiety. Sedentary and leisure screen time were positively correlated with negative affect. Our findings suggest that maintaining positive affect, engaging in PA, and limiting leisure screen time may be important for child mental health during stressful periods.

**Statement of Relevance:** There is increasing concern regarding how the COVID-19 pandemic may impact the psychological and physical health of children. To date, studies on mental health during the pandemic in children are limited. We investigated links between activity levels and psychological outcomes in children during the height of the “stay-at-home” measures. We found that children had anxiety scores that were more than 5 standard deviations greater than normative values from healthy pediatric populations prior to the pandemic, and 94% of children exceeded the American Academy of Pediatrics recommendations on leisure screen time. Positive affect and physical activity were associated with reduced anxiety levels in children during the pandemic. These findings highlight the important mental health benefits of maintaining positive affect, engaging in physical activity, and limiting leisure screen time for children, especially during stressful periods.

## Introduction

The coronavirus disease 2019 (COVID-19) was declared a national public health emergency in the United States in March 2020. In order to prevent the spread of the virus, US state and local governments implemented unprecedented “stay-at-home” orders starting in mid-March, 2020, including closures of primary and secondary schools across the nation (https://plus.google.com/+UNESCO, 2020). Consequently, children’s access to social support resources and opportunities for physical activity may have been limited during home confinement.

In addition to the high COVID-19 death toll, which surpassed 200,000 in the U.S. by the end of September 2020 (“Mortality Analyses”), there is increasing concern regarding the potential collateral damage to physical and mental health during the COVID-19 pandemic. Over one-third of American adults reported that the pandemic is having a serious impact on their mental health (“New Poll: COVID-19 Impacting Mental Well-Being: Americans Feeling Anxious, Especially for Loved Ones; Older Adults Are Less Anxious,”), and a recent study among US adults demonstrated that the prevalence of symptoms of psychological distress were significantly higher during the pandemic compared to two years earlier (McGinty, Presskreischer, Han, & Barry, 2020). While little is known about the impact of the pandemic on the mental health of US children, school-aged children in China reported experiencing depressive symptoms during their nationwide lockdown (Xie et al., 2020). Furthermore, other recent studies in China, Italy, and the United States have also shown that children are engaging in less physical activity and increased sedentary behavior and screen time during the pandemic (Dunton, Do, & Wang, 2020; Pietrobelli et al., 2020; M. Xiang, Zhang, & Kuwahara, 2020). Notably, poor mental health outcomes have been identified as independent risk factors for several chronic conditions such as obesity, diabetes, and cardiometabolic disease (Golden et al., 2008; Rudisch & Nemeroff, 2003); additionally, low physical activity levels, heightened screen time, and increased sedentary behavior are also associated with these conditions (Byun, Dowda, & Pate, 2012; Shrestha & Copenhaver, 2015). Given that health behavior trajectories in childhood are likely to endure through adulthood (see review by Shrestha and Copenhaver (2015)), these findings raise alarming public health implications.

Correspondingly, studies conducted prior to the pandemic demonstrated a clear link between psychological well-being and lifestyle behaviors. Prior studies have shown that positive affect, defined as the tendency of an individual to experience positive emotions, such as enthusiasm and joy (*Positive Emotion*, 2014), is associated with increased engagement in exercise (Pasco et al., 2011; Peterson et al., 2012). Positive affect is negatively correlated with symptoms and diagnoses of anxiety and depression (Peterson et al., 2012; Watson, Clark, & Carey, 1988), and multiple reports have established that physical activity reduces symptoms of anxiety among children (Kiluk, Weden, & Culotta, 2009; Parfitt & Eston, 2005). Conversely, studies have shown that during childhood and adolescence, both sedentary behavior and screen time are associated with increased risk for depressive symptoms and negative affect, which refers to the tendency to experience more intense negative emotions (García-Hermoso, Hormazábal-Aguayo, Fernández-Vergara, Olivares, & Oriol-Granado, 2020; Sund, Larsson, & Wichstrøm, 2011).

While COVID-19 restrictions may impact both the psychological and physical health of children, to our knowledge, no study to date has examined how children’s positive and negative affect, physical activity and sedentary behaviors relate to their anxiety levels. The aim of the current study was to assess children’s positive and negative affect and activity levels during the COVID-19 “stay-at-home” order, and to examine relationships between affect, anxiety, physical activity and sedentary behaviors in children.

## Methods

### Participants

Participants in this study were recruited from the existing BrainChild study, an observational study that includes healthy, typically developing children who were recruited at ages 7 to 11 years old during years 2014-2018 for entry into the study and followed with annual visits. The studies reported here were completed between one and four years after initial entry into the study. Children in the BrainChild cohort were born at a Kaiser Permanente Southern California (KPSC) Hospital and had no history of psychiatric, neurological, or other significant medical disorders.

Due to the COVID-19 pandemic, participants were unable to complete in-person consents to participate in this sub-study. Therefore, an amendment to the original Institutional Review Board (IRB) protocol was sent to the IRBs at the University of Southern California (#HS-15-00540) and KPSC (#10282) for virtual follow-up visits. Both USC and KPSC IRBs approved this sub-study if participants had a recent in-person follow-up visit that occurred within a year. During the prior in-person follow-up visit, participants’ parents gave written informed consent, and children provided written informed assent to participate in longitudinal studies. Additionally, participants gave verbal informed consent prior to participating in the phone or video interviews. Of the 162 participants enrolled in the larger BrainChild Study, 82 participants had recent 1-year follow-up visits and, therefore, were eligible to participate in this ancillary study. During in-person visits that had occurred within one year of the virtual visits, height (cm) to the nearest 0.1 cm was collected using a stadiometer and weight (kg) was collected using a calibrated digital scale. BMI was calculated using the standard formula, weight (kg) divided by height (m^2^). BMI z-scores and BMI percentiles (age and sex-specific standard deviation scores) were determined based on Center for Disease Control (CDC) standards (*About Child & Teen BMI* | *Healthy Weight* | *CDC*, 2018).

### Exposure

All of the children were residents of California, which was under a statewide “stay-at-home” lockdown starting March 19, 2020. Data collection took place during Phase 1/Phase 2 of the order from April 22^th^-July 29^th^, 2020, wherein all schools were closed for in-person instruction from April 22 – May 29, 2020.

### Phone Visit

Trained staff members contacted participants’ parents from the existing BrainChild cohort. The study included two phone or video call visits with both the participant and a parent present. Each visit occurred on average 34 days apart ranging from 27 to 73 days, interquartile range 30 – 35 days. Visit one occurred from April 22^nd^ to June 26^th^, 2020. Visit two occurred from May 22^nd^ to July 29^th^, 2020. All questionnaires were read aloud to each participant by the trained staff member, and then the participant gave their answers verbally.

### Physical Activity Assessment

At each phone call visit, physical activity was assessed using a 24-h physical activity recall (PAR) (Ainsworth et al., 2011; Weston, Petosa, & Pate, 1997). A trained staff member asked participants, with a parent present to offer input, to recall all of their activities from 7:00am to 12:00am in 30-minute blocks for the day prior. The activities were recorded and classified based on a 73-item reference list. The participant was also asked to rate the intensity of each activity as either “Light” (slow breathing, little/no movement), “Moderate” (normal breathing and some movement), “Hard” (increased breathing and moderate movement), or “Very Hard” (hard breathing and quick movement). Each activity was then categorized as either sedentary, moderate-to-vigorous physical activity (MVPA), or vigorous physical activity (VPA), with their associated metabolic equivalent (MET) values obtained from the Compendium of Physical Activities (Ainsworth et al., 2011). Activities with MET values >1 and ≤1.5 were classified as sedentary, METs ≥3 as MVPA, and METs ≥6 as VPA. Sleep blocks were classified as METs=1.0. Examples of physical activity classifications include: walking (MVPA) or swimming laps (VPA). Examples of sedentary activities include, reading a book, sitting in class and as well as any sedentary screen time activities. In addition, leisure screen time (MET=1.5 per 30-minute block) was obtained by adding time spent in the following leisure activities: watching TV or movies, playing video games, and surfing the internet while sedentary.

### State-Trait Anxiety Inventory for Children

At each phone call visit, state anxiety was assessed via the State-Trait Anxiety Inventory for Children (STAIC). Only items for the state-anxiety (S-Anxiety) scale were completed, given that the research question was concerned with how children were acutely responding to the pandemic, and by design, S-Anxiety scores are influenced by the child’s immediate environment (Spielberger & Edwards, 1973). The STAIC S-Anxiety scale is composed of 20 statements, and children are instructed to base their answers on how they feel *at that particular moment*. Each STAIC S-Anxiety item is a 3-point rating scale with a stem of “I feel”; half of the items are reflective of the presence of anxiety (i.e. nervous, worried), while the other half are indicative of the absence of anxiety (i.e. calm, pleasant) (Spielberger & Edwards, 1973). Values of 1, 2, or 3 are assigned for each of the three answer choices, and scores range from 20-60, with higher scores representing higher state anxiety.

### Positive and Negative Affect Schedule for Children

At each phone call visit, both positive and negative affect were assessed using the Positive and Negative Affect Schedule for Children (PANAS-C). The current study utilized the shortened 10-item PANAS-C, which includes a 5-item positive affect scale (*joyful, cheerful, happy, lively, proud*) and a 5-item negative affect scale (*miserable, mad, afraid, scared, sad*), with a 5-point Likert scale ranging from 1 (“very slightly or not at all) to 5 (“extremely”) (Ebesutani et al., 2012). The participants were instructed to answer each item reflecting to what extent they have felt this way *during the past few* weeks in order to capture a wider temporal range of affect during the pandemic. Scores range from 5-25 for each affect subscale, with higher scores representing higher affect.

### Statistical Analysis

To minimize data reporting errors, measures taken from each of the two visits were averaged and used for all analyses. Descriptive statistics including mean±SD, median (IQR), ranges and frequencies were reported. Correlations and linear regression models were used to test associations between emotional regulatory processes (positive affect and negative affect), physical activity (MVPA and VPA) and sedentary behaviors (leisure screen time, sedentary time) with state anxiety. We assessed whether emotional regulatory processes and physical activity were independently associated with state anxiety by including both in the same model. Covariates included in each linear regression model were child age, sex, socioeconomic status (SES) and BMI z-score because these are factors known to influence mental health and physical activity levels (Belcher et al., 2010; Fakhouri, Hughes, Brody, Kit, & Ogden, 2013; Mason et al., 2019; Nader, Bradley, Houts, McRitchie, & O’Brien, 2008; Topçu, Orhon, Tayfun, Uçaktürk, & Demirel, 2016; Zhu et al., 2019). SES was assessed using household income at birth, estimated based on census tract of residence and expressed as a continuous variable, and maternal education at birth, extracted from birth certificates in the electronic medical record as a categorical variable with the following categories: “high-school or some high-school”, “some college” and “college and post-education” (A. H. Xiang et al., 2015). Time spent in MVPA and VPA were not normally distributed, and a square-root transformation was applied to normalize the distribution prior to regression analyses. P-values <0.05 were interpreted as statistically significant. SAS 9.4 statistical software (SAS Institute, Cary, NC USA) was used for all data analyses.

## Results

Of the 82 participants from the BrainChild study who had completed at least one longitudinal follow-up visit, 65 participants completed one phone-call or video visits during the state mandated “stay-at-home” order, and 64 of these participants completed a second phone-call or video visit approximately one month later. Child age ranged from 9 to 15 years and the mean ±SD age was 11.84 ± 1.28 years, 63% of participants were female, and 53% were of a healthy-weight defined as a BMI percentile less than 85. Children’s mean ±SD positive affect scores were 16.16 ± 4.10, negative affect scores were 8.27 ± 3.15, and STAIC S-Anxiety scores were 47.37 ± 3.15 **(Table 1)**. Mean ±SD scores for other healthy pediatric samples of a similar age range are as follows: positive affect (17.34 ± 3.07) (Bauer et al., 2019; Zink et al., 2020), negative affect (7.20 ± 2.44) (Bauer et al., 2019; Zink et al., 2020), state anxiety (29.78 ± 1.17) (Mestre et al., 2019; Spielberger & Edwards, 1973; Zink et al., 2020). Thus, compared to healthy pediatric studies conducted prior to the pandemic, state anxiety in our cohort was five standard deviations greater. Mean ±SD BMI z-score was 0.87 ± 1.15, the mean sedentary time was 677.81 ± 110.06 (min/day), mean leisure screen time was 376.64 ± 171.70 (min/day), median (IQR) MVPA was 45 (67.5) (min/day), and median VPA was 0 (15) (min/day). Forty-seven children (73%) did not engage in any VPA. Additionally, 94% (60/64) of children reported more than 2 hours of leisure screen time per a day, exceeding the American Academy of Pediatrics recommended 2 hour limit for screen time (Fakhouri et al., 2013). The mean sedentary time reported by the US National Health and Nutrition Examination Survey for similarly aged children was 498.20 ± 162.20 (min/day), screen time was 264.80 ± 152.90 (min/day), MVPA was 40.40 ± 26.50 (min/day) and VPA was 7.60 ± 10.02 (min/day) (Belcher et al., 2010; Hunt et al., 2019).

**Table 1.** Participant Demographics (N=64).

| **Variable** | **N (%) or Mean±SD or Median (IQR)** | **Range** |
| --- | --- | --- |
| **Age (years)** | 11.84±1.28 | 9.36 to 15.42 |
| **Sex** | Girls: 40 (63%)<br>Boys: 24 (37%) |  |
| **BMI z-score** | 0.87±1.15 | -1.44 to 2.65 |
| **Child BMI Percentile Category** | Healthy-weight: 34 (53%)<br>Overweight: 10 (16%)<br>Obese: 20 (31%) |  |
| **STAIC-State Anxiety** | 47.37±3.15 | 40.50 to 52 |
| **PANAS for children-Positive** | 16.16±4.10 | 7 to 25.00 |
| **PANAS for children-Negative** | 8.27±2.93 | 5 to 19.50 |
| **Sedentary time (min)** | 677.81±110.06 | 390 to 900.00 |
| **Screen time (min)** | 376.64±171.70 | 105 to 750.00 |
| **Time in MVPA (min)** | 45 (67.5) | 0 to 240.00 |
| **Time in VPA (min)** | 0 (15) | 0 to 150.00 |
| **Children who engaged in VPA** | VPA: 17 (27%)<br>No VPA: 47 (73%) |  |
| **Maternal Education** | <=High school: 6 (9%)<br>Some college: 19 (30%)<br>College and post: 39 (61%) |  |
| **<sup>a</sup>Household Income** | 0<=income <30 000: 5 (8%)<br>30000<=income <50 000: 20 (31%)<br>50000<=income <70 000: 23 (36%)<br>70000<=income <90 000: 8 (13%)<br>90000>=income: 8 (13%) |  |

<sup>a</sup>Some percentages may not be equal to 100% due to rounding.
Abbreviations: BMI, body mass index. MVPA, moderate-to-vigorous physical activity. VPA, vigorous physical activity. STAIC, State-Trait Anxiety Inventory for Children (state anxiety). PANAS, Positive and Negative Affect Schedule for Children (positive and negative affect).

For the relationship between emotional regulatory and anxiety measures **(Table 2)**, positive affect was associated with lower state anxiety in unadjusted models (correlation coefficient r=-0.50, p<0.001; regression β=-0.38, 95% CI: −0.55,−0.22, p<0.001) and after adjusting for child age, sex, SES, and BMI z-score (β=−0.40, 95% CI: −0.57, −0.22, p<0.001). However, negative affect was not associated with state anxiety in unadjusted models (r=−0.03, p=0.78; β=−0.04, 95% CI: −0.31, 0.23, p=0.79) or in models adjusted for child age, sex, SES and BMI z-score (β=−0.004, 95% CI: −0.28, 0.28, p=0.98).

**Table 2.** Association between Affect and State Anxiety.

| **Predictor Variables** | **Beta (95% CI)** | **p-value** | **Covariates** |
| --- | --- | --- | --- |
| **Positive Affect** | -0.38 (-0.55, -0.22) | <0.001*** | Unadjusted |
|  | -0.40 (-0.57, -0.22) | <0.001*** | Age, sex, SES and BMI z-score |
|  | -0.36 (-0.53, -0.18) | <0.001*** | Age, sex, SES, BMI z-score and MVPA |
| **Negative Affect** | -0.04 (-0.31, 0.23) | 0.79 | Unadjusted |
|  | -0.004 (-0.28, 0.28) | 0.98 | Age, sex, SES and BMI z-score |
|  | -0.05 (-0.32, 0.22) | 0.72 | Age, sex, SES, BMI z-score and MVPA |

\*\*\*p-value<0.001
Abbreviations: SES, socioeconomic status (family income and maternal education). MVPA, moderate-vigorous physical activity.

For the relationship between physical activity and anxiety measures (**Table 3**), MVPA was associated with less state anxiety in both the unadjusted model (r=−0.34, p=0.007; β=−0.27, 95% CI: −0.47, −0.08, p=0.007), and after adjusting for child age, sex, SES and BMI z-score (β=−0.27, 95% CI: −0.47,−0.06, p=0.01). VPA was not associated with lower state anxiety in either the unadjusted (r=−0.08, p=0.52; β=−0.11, 95% CI: −0.46, 0.23, p=0.52) or adjusted models (β=−0.07, 95% CI: −0.42, 0.29, p=0.72). Sedentary time was also not associated with state anxiety before (r=0.06, p=0.63; β=0.002, 95% CI: −0.01, 0.01, p=0.63) and after adjusting for covariates (β=0.003, 95% CI: −0.004, 0.01, p=0.44) (**Table 3)**. Leisure screen time was not associated with state anxiety in the unadjusted (r=0.10, p=0.42; β=0.002, 95% CI: −0.003, 0.01, p=0.42) and adjusted models (β=0.003, 95% CI: −0.002, 0.01, p=0.24) (**Table 3)**.

**Table 3.** Association between Activity Levels and State Anxiety.

| **Predictor Variables** | **Beta (95% CI)** | **p-value** | **Covariates** |
| --- | --- | --- | --- |
| **Moderate-to-Vigorous Physical Activity** | -0.27 (-0.47, -0.08) | 0.007** | Unadjusted |
|  | -0.27 (-0.47, -0.06) | 0.01* | Age, sex, SES and BMI z-score |
|  | -0.20 (-0.38, -0.01) | 0.04* | Age, sex, SES, BMI z-score and positive affect |
| **Vigorous Physical Activity** | -0.11 (-0.46, 0.23) | 0.52 | Unadjusted |
|  | -0.07 (-0.42, -0.29) | 0.72 | Age, sex, SES and BMI z-score |
|  | -0.04 (-0.35, 0.28) | 0.83 | Age, sex, SES, BMI z-score and positive affect |
| **Sedentary Time** | 0.002 (-0.005, 0.01) | 0.63 | Unadjusted |
|  | 0.003 (-0.004, 0.01) | 0.44 | Age, sex, SES and BMI z-score |
|  | 0.001 (-0.01, 0.01) | 0.73 | Age, sex, SES, BMI z-score and positive affect |
| **Screen Time** | 0.002 (-0.003, 0.01) | 0.42 | Unadjusted |
|  | 0.003 (-0.002, 0.01) | 0.24 | Age, sex, SES and BMI z-score |
|  | 0.001 (-0.003, 0.01) | 0.54 | Age, sex, SES, BMI z-score and positive affect |

\*\*p-value<0.01
\*p-value<0.05
Abbreviations: SES, socioeconomic status (family income and maternal education)

Analyses assessing the independent associations between affect score and physical activity with state anxiety showed that both remained associated with state anxiety when adjusting for each other. Positive affect remained associated with lower state anxiety after adjusting for MVPA (β changed from −0.40 to −0.36, 95% CI: −0.54, −0.18, p<0.001, **Table 2**). MVPA also remained associated with lower state anxiety after further adjusting for positive affect (β changed from −0.27 to −0.20, 95% CI: −0.38, −0.01, p=0.04, **Table 3**).

When examining relationships between emotional regulatory responses (i.e., positive and negative affect) and physical activity measures, we found that negative affect was correlated with sedentary time (r=0.28, p=0.02) and leisure screen time (r=0.40, p=0.001) (**Table 4)**. After adjusting for child age, sex, SES and BMI z-score, negative affect remained correlated with leisure screen time (r=0.40, p=0.002) and sedentary time (r=0.28, p=0.03) (**Table 5**). Positive affect was not related to any of the physical activity measures (**Tables 5 and 6**).

**Table 4.** Pearson R correlations between emotional regulatory responses (positive and negative affect), state anxiety, physical activity (MVPA and VPA), screen time, sedentary behavior and child adiposity in unadjusted model.

|  | **Positive Affect** | **Negative Affect** | **State Anxiety** | **MVPA** | **VPA** | **Screen Time** | **Sedentary Time** | **BMI z-score** |
| --- | --- | --- | --- | --- | --- | --- | --- | --- |
| **Positive Affect** | 1 | -0.16 | **-0.50\*\*** | 0.10 | 0.07 | -0.11 | -0.17 | -0.03 |
| **Negative Affect** | -0.16 | 1 | -0.03 | -0.08 | -0.19 | **0.40\*\*** | **0.28\*** | 0.05 |
| **State Anxiety** | **-0.50\*\*** | -0.03 | 1 | **-0.34\*** | -0.08 | 0.10 | 0.06 | -0.09 |
| **MVPA** | 0.10 | -0.07 | **-0.34\*** | 1 | **0.48\*\*** | -0.16 | **-0.41\*\*** | -0.06 |
| **VPA** | 0.07 | -0.19 | -0.08 | **0.48\*\*** | 1 | -0.13 | -0.13 | -0.01 |
| **Screen Time** | -0.17 | **0.40\*\*** | 0.10 | -0.16 | -0.13 | 1 | **0.59\*\*** | 0.05 |
| **Sedentary Time** | -0.11 | **0.28\*** | 0.06 | **-0.41\*\*** | -0.13 | **0.59\*\*** | 1 | -0.08 |
| **BMI z-score** | -0.03 | 0.05 | -0.09 | -0.06 | -0.01 | 0.05 | -0.08 | 1 |

\*Denotes p-value<0.05.
\*\*Denotes p-value<0.01.
MVPA=moderate-to-vigorous physical activity. VPA=vigorous physical activity. MVPA and VPA square-root transformed.

**Table 5.** Pearson R correlations between emotional regulatory states (positive and negative affect), state Anxiety, physical activity (MVPA and VPA), screen time, sedentary behavior, and child adiposity in adjusted models.

|  | **Positive Affect** | **Negative Affect** | **State Anxiety** | **MVPA** | **VPA** | **Screen Time** | **Sedentary Time** | **BMI z-score** |
| --- | --- | --- | --- | --- | --- | --- | --- | --- |
| **Positive Affect** | 1 | -0.16 | **-0.48\*\*** | 0.16 | 0.03 | -0.17 | -0.13 | -0.09 |
| **Negative Affect** | -0.16 | 1 | -0.01 | -0.12 | -0.18 | **0.40\*\*** | **0.28\*** | 0.05 |
| **State Anxiety** | **-0.48\*\*** | -0.01 | 1 | **-0.33\*** | -0.05 | 0.16 | 0.10 | -0.04 |
| **MVPA** | 0.16 | -0.12 | **-0.33\*** | 1 | **0.50\*\*** | **-0.26\*** | **-0.50\*\*** | 0.05 |
| **VPA** | 0.03 | -0.18 | -0.05 | **0.50\*\*** | 1 | -0.16 | -0.16 | -0.04 |
| **Screen Time** | -0.17 | **0.40\*** | 0.16 | **-0.26\*** | -0.16 | 1 | **0.58\*\*** | 0.03 |
| **Sedentary Time** | -0.13 | **0.28\*** | 0.10 | **-0.50\*\*** | -0.16 | **0.58\*\*** | 1 | -0.10 |
| **BMI z-score** | -0.09 | 0.05 | -0.04 | 0.05 | -0.04 | 0.03 | -0.10 | 1 |

Models adjusted for child age, sex and socioeconomic status and BMI z-score.
\*Denotes p-value<0.05.
\*\*Denotes p-value<0.01.
MVPA=moderate-to-vigorous physical activity. VPA=vigorous physical activity. MVPA and VPA square-root transformed.

## Discussion

We provide the first results from the United States that examined how emotional regulatory responses, measured from positive and negative affect scores, related to anxiety levels and physical activity levels of children during the pandemic. In California, the “stay-at-home” orders began in late March and were lifted at the end of May. During this time, we collected questionnaires on affect, state anxiety, physical activity and sedentary behaviors during the peak of the “stay-at-home” order to infer not only how anxiety levels were impacted, but also the role of positive and negative affect and physical activity levels on anxiety levels in children. We found that state anxiety levels of children in our cohort during the “stay-at-home” order were more than five standard deviations greater than the mean reported by other healthy pediatric populations prior to the pandemic (Mestre et al., 2019; Spielberger & Edwards, 1973; Zink et al., 2020). Additionally, children reported greater screen and sedentary time than similarly aged children from the NHANES (Belcher et al., 2010; Hunt et al., 2019). Interestingly, we found that positive affect, negative affect and child physical activity levels in our cohort were similar to those reported by other pediatric studies conducted prior to the pandemic (Bauer et al., 2019; Belcher et al., 2010; Hunt et al., 2019; Zink et al., 2020).

Prior studies in countries first struck by the pandemic noted an increase in symptoms of depression and anxiety in both children and adults (Cao et al., 2020; Xie et al., 2020; Zhang, Wang, Rauch, & Wei, 2020). However, none of these studies have investigated relationships between affect and mental health during the pandemic. Our study showed that positive affect was related to lower state anxiety levels in children, independent of age, sex, socioeconomic status, physical activity, and BMI. These findings are in keeping with larger cross-sectional studies suggesting that positive affect is protective against anxiety during stressful times (O’Hara, Armeli, Boynton, & Tennen, 2014; Sewart et al., 2019). Therefore, promoting methods to maintain positive affect, such as educational interventions that encourage practicing gratitude (Froh et al., 2014) and mindfulness (Kang et al., 2018), may be beneficial to children during times of heightened stress, such as the COVID-19 pandemic. However, future studies are needed to test this possibility.

While anxiety levels have been documented to be increased in adult populations during the pandemic (Cao et al., 2020; McGinty et al., 2020; Zhang et al., 2020), the impact of COVID-19 restrictions on child mental health is sparse (Xie et al., 2020). Similar to the one study published in children during the COVID-19 lockdown in China (Xie et al., 2020), we found that children during the “stay-at-home” order reported greater state anxiety compared to other pediatric samples prior to the pandemic (Mestre et al., 2019; Spielberger & Edwards, 1973; Zink et al., 2020). In addition to higher anxiety symptoms, Xie et al. found that school-aged children in China reported higher depression symptoms (Xie et al., 2020). The authors hypothesized that a reduction in outdoor activities and social interactions may have contributed to increased depression and anxiety symptoms. Correspondingly, several studies have found that engaging in physical activity is beneficial for mental health (Crews, Lochbaum, & Landers, 2004; Parfitt & Eston, 2005; Parfitt, Pavey, & Rowlands, 2009). Interestingly, we found that children who engaged in more physical activity had less reported state anxiety, independent of age, sex, socioeconomic status, positive affect, and BMI. Taken together, these findings support a critical role for physical activity in promoting the health and well-being of children. Engagement in physical activity may be particularly important for children to help reduce anxiety during stressful periods.

Similar to other studies in children during the “stay-at-home” order, we found an increase in both sedentary and leisure screen time compared to nationally representative pediatric samples before the “stay-at-home” order (Dunton et al., 2020; Fakhouri et al., 2013; Pietrobelli et al., 2020; M. Xiang et al., 2020). Prior to the pandemic, the American Academy of Pediatrics recommended that children engage in less than 2 hours a day of leisure screen time (Fakhouri et al., 2013). However, children in our cohort reported an average of 6 hours a day of leisure screen time. Additionally, children reported spending 11 hours a day being sedentary and in leisure screen time. Importantly, prior studies in youth have shown that excessive screen and sedentary time is associated with increased depressive symptoms and negative affect (García-Hermoso et al., 2020; Sund et al., 2011). Correspondingly, we found that increased leisure screen time was associated with negative affect in children, independent of age, sex, socioeconomic status, and BMI. Similarly, more time spent sedentary was also associated with negative affect. While our study design does not allow us to determine the directionality of the relationship between leisure screen time and negative affect, our findings are in concert with previous large cross-sectional studies demonstrating a dose-dependent relationship between screen-based activities and depressive symptomatology such as negative affect in children (García-Hermoso et al., 2020; Yang, Helgason, Sigfusdottir, & Kristjansson, 2013). Therefore, future studies should consider investigating if limiting excessive leisure screen time could reduce the risk for negative affect among children.

Our study was able to collect repeated measures of affect, anxiety and behavioral health questionnaires in children over two months during the peak of the “stay-at-home” orders, but we did not have baseline measures of affect or anxiety in this cohort prior to the pandemic to compare to the measures collected during the pandemic. While we did compare affect, anxiety, and activity levels in our cohort to other healthy pediatric populations prior to the pandemic, it is worth noting that the normative comparisons that we used for anxiety were either limited in sample size (Mestre et al., 2019; Zink et al., 2020) or not recent (Spielberger & Edwards, 1973). However, to the best of our knowledge, there are no recent and large sample size normative STAIC state-anxiety comparisons available in US children and/or adolescents. Moreover, we assessed a limited number of behavioral factors that predicted levels of anxiety among children during COVID-19 restrictions. Future pandemic-related studies should consider assessing other potential environmental and psychosocial risk and protective factors for anxiety in children, such as sleep (Alfano, Ginsburg, & Kingery, 2007) and social support (Sandler, Miller, Short, & Wolchik, 1989). Additionally, we used a self-reported physical activity recall which is subject to participant bias. However, self-report recalls provided pertinent information about specific physical activities engaged in during the “stay-at-home” order. Finally, our small sample size may limit the generalizability of our findings.

## Conclusions

Overall, this study found that children reported heightened anxiety during the COVID-19 “stay-at-home” orders when compared to normative values from pediatric populations prior to the pandemic. Importantly, we found that children who reported greater positive affect reported lower anxiety symptoms. We also found that children who engaged in more physical activity reported lower anxiety symptoms and this relationship was independent of positive affect. Additionally, negative affect was positively correlated with both sedentary time and leisure screen time. These study findings highlight the important mental health benefits of maintaining positive affect, engaging in physical activity, and limiting leisure screen time, especially during stressful periods.

## Data Availability

Data availability: The datasets generated during and analyzed during the current study are available from the corresponding author (K.A.P.), on reasonable request.

## Data availability

The datasets generated during and analyzed during the current study are available from the corresponding author (K.A.P.), on reasonable request.

## Abbreviations

COVID-19: coronavirus disease 2019
STAIC: State-Trait Anxiety Inventory for Children
PANAS-C: Positive and Negative Affect Schedule for Children
MET: metabolic equivalent
MVPA: moderate-to-vigorous physical activity
VPA: vigorous physical activity
BMI: body mass index
NHANES: National Health and Nutrition Examination Survey

## Contributors’ Statement

Dr. Alves performed statistical analyses, drafted the initial manuscript, and reviewed and revised the manuscript. Ms. Yunker collected and organized data, drafted the initial manuscript, and reviewed and revised the manuscript. Ms. DeFendis collected and organized data. Dr. Xiang contributed to study concept and design, obtained funding and provided study supervision, performed statistical analyses, drafted the initial manuscript, and reviewed and revised the manuscript. Dr. Page contributed to study concept and design, obtained funding and provided study supervision, drafted the initial manuscript, and reviewed and revised the manuscript. All authors critically reviewed the manuscript for important intellectual content, approved the final manuscript as submitted, and agree to be accountable for all aspects of the work.

## Acknowledgments

The authors would like to thank the families who participate in the BrainChild Study. The authors would also like to thank Ana Romero for managing the BrainChild study, Mayra Martinez and Janet Mora-Marquez for recruiting volunteers and helping collect participant data.

